# A study of the clearness rate of fetal Sylvian fissure shown on different sections by prenatal ultrasound

**DOI:** 10.1101/2021.04.21.21255847

**Authors:** Xu Pingping, Zhang Dirong, Shi Yu, Kong Fengbei, Yao Chunxiao, She Ying, Wu Guoru

## Abstract

**Objective:** To compare the clearness rate of fetal Sylvian fissure revealed by prenatal ultrasonography on trans-cerebellar section and trans-thalamic section, and to provide scientific basis for selecting the best section of prenatal ultrasound to evaluate the developmental parameters related to fetal Sylvian fissure.

**Methods:** This was a retrospective study. We analyzed all the fetal images on trans-cerebellar section and trans-thalamic section stored in the imaging system who were undergoing grade III prenatal ultrasound examination in our hospital from January 2017 to December 2018. The gestational age was divided into three groups which were 21 to 24 weeks, 25 to 28 weeks and 29 to 32 weeks respectively. The evaluation criteria for the clear appearance of the Sylvian fissure were complete morphology of the Sylvian fissure and clear apical and anteriorly edges of the Sylvian fissure. The results of showing Sylvian fissure clearly were divided into four groups: only on trans-cerebellar section shown clear, only on trans-thalamic section shown clear, both on trans-cerebellar section and trans-thalamic section shown clear, and both on trans-cerebellar section and trans-thalamic section not shown clear. The clearness rate of each group and the total clearness rate of each section were analyzed statistically.

**Results:** The clearness rates of showing Sylvian fissure only on trans-cerebellar section were 62.3% at 21 to 24 weeks, 70.1% at 25 to 28 weeks and 63.6% at 29 to 32 weeks. The clearness rates of showing Sylvian fissure only on trans-thalamic section were 11.1% at 21 ∼ 24 weeks, 10.5% at 25 ∼ 28 weeks and 9.1% at 29 ∼ 32 weeks. The clearness rates of showing Sylvian fissure both on trans-cerebellar and trans-thalamic sections were 22.1% at 21 to 24 weeks, 15.0% at 25 to 28 weeks and 13.0% at 29 to 32 weeks. The unclearness rates of showing Sylvian fissure both on trans-cerebellar and trans-thalamic sections were 4.5%at 21 ∼ 24 weeks, 4.4% at 25 ∼ 28 weeks and 14.3% at 29 ∼ 32 weeks. The clearness rate of showing Sylvian fissure on trans-cerebellar section was significantly higher than on trans-thalamic section (P < 0.05). The total clearness rate of showing Sylvian fissure on trans-cerebellar section and trans-thalamic section were 83.1% and 28.3% respectively.

**Conclusion:** In this study, it was suggested that prenatal ultrasound selection of trans-cerebellar section is better than trans-thalamic section in evaluating the developmental parameters related to the Sylvian fissure.

## INTRODUCTION

Fetal cortical dysplasia could lead to postnatal mental retardation, motor disorders and intractable epilepsy^1^, so prenatal diagnosis of cortical dysplasia is particularly important. However, cortical developmental disorder is a difficult problem in prenatal diagnosis^2^. The deformity of cerebral cortical development was mainly manifested as delayed sulci development^3^. The Sylvian fissure is one of the main cerebral sulcus and its development is representative. At present, there have been some studies on the development rules of normal fetal Sylvian fissure, but the research sections are not uniform, mainly including trans-thalamic section and some other sections, and the research conclusions differ greatly^4-8^, so there is no research result for promotion and application at present. In our previous work, we found that the trans-cerebellar section showed the Sylvian fissure more clearly than the trans-thalamic section. Theoretically, the ultrasonic beam on the transverse section of cerebellar was more perpendicular to the Sylvian fissure, so we hypothesized that the trans-cerebellar section should also show the lateral fissure more clearly. In order to further confirm our hypothesis, we retrospectively analyzed the images saved in the workstation during our earlier screening of fetal structural malformations, and compared the clearness rate of fetal Sylvian fissure shown on trans-cerebellar section and trans-thalamic section, so as to provide a scientific basis for selecting the best section for prenatal ultrasound to evaluate developmental parameters related to Sylvian fissure.

## METHODS

This was a retrospective study. We analyzed all the fetal images on trans-cerebellar section and trans-thalamic section stored in the imaging system who were undergoing grade III prenatal ultrasound examination in our hospital from January 2017 to December 2018. There was no selection bias in the subjects. To ensure the authenticity and reliability of the data, the inclusion criteria were as follows: normal fetus without structural malformation, the stored images of each fetus in the imaging system included trans-thalamic section and trans-cerebellar section, low-risk pregnancies including without diabetes, hypertension and other pregnancy risk factors. Exclusion criteria were as follows: fetuses with known malformations or chromosomal abnormalities, multiple pregnancies, the images with trans-thalamic or/and trans-cerebellar section not standard; no neurological symptoms within 6 months after birth. This study was approved by the Peking University Shenzhen Hospital and Health Service Human Research Ethics Committee. All pregnant women gave informed consent to this study. The pregnant women ranged in age from 20 to 41 years old, with an average age of 28 years old. The gestational age ranged from 21 to 32 weeks, with an average gestational age of 26 weeks.

The ultrasound machine used in this study was a Voluson E8 or E10 (GE Healthcare Ultrasound) with a 4-8MHz transabdominal 2D transducer. The gestational age was divided into three groups which were 21 to 24 weeks, 25 to 28 weeks and 29 to 32 weeks respectively. The evaluation criteria for the clear appearance of the Sylvian fissure was complete morphology of the Sylvian fissure and clear apical and anteriorly edges of the Sylvian fissure. The results of showing Sylvian fissure clearly were divided into four groups: only on trans-cerebellar section shown clear, only on trans-thalamic section shown clear, both on trans-cerebellar section and trans-thalamic section shown clear, and both on trans-cerebellar section and trans-thalamic section not shown clear. The clearness rate of each group and the total clearness rate of each section were analyzed statistically.

SPSS 26.0 statistical software was used to analyze and process the data. The main steps were as follows:(1) the sample size of all gestational weeks was counted, and the sample size and clearness rate of each group of showing Sylvian fissure were counted according to gestational age, (2) the sample size and clearness rate of Sylvian fissure could be clearly shown on different sections of each gestational age group were counted respectively; (3) the total clearness rate of Sylvian fissure in different sections was counted; (4) the counting data were expressed as n /%, and the comparison of the clearness rate of the Sylvian fissure of the fetus in each section was tested by chi-square test. P < 0.05 was considered statistically significant.

## RESULTS

A total of 6980 subjects were included in the study. The sample size and percentage distribution of gestational weeks in each group were as follows: 3182 cases (45.6%) at 21 to 24 weeks, 2456 cases (35.2%) in at 25 to 28 weeks, and 1342 cases (19.2%) at 29 to 32 weeks. The data was shown in Table 1.

**Table 1.**
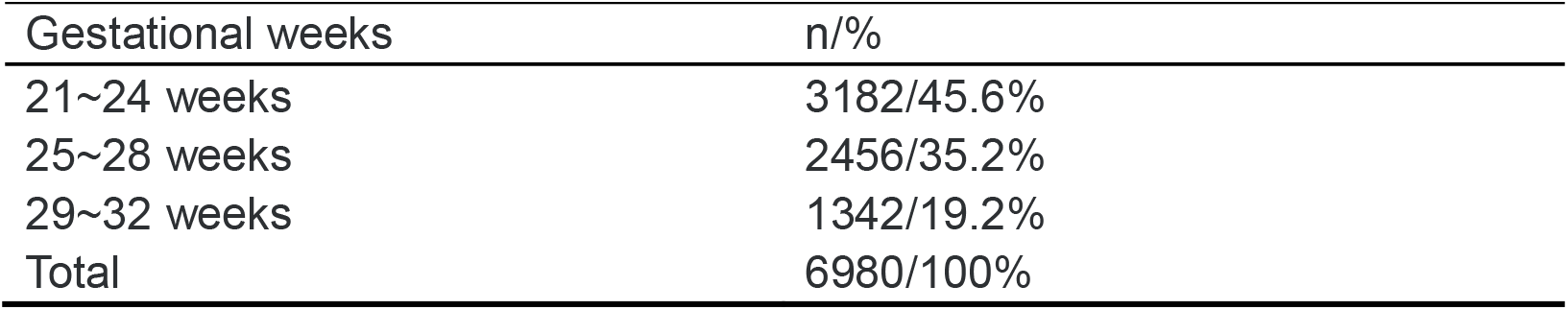
Sample size and percentage of each gestational age group (n/%)

| Gestational weeks | n/% |
| --- | --- |
| 21~24 weeks | 3182/45.6% |
| 25~28 weeks | 2456/35.2% |
| 29~32 weeks | 1342/19.2% |
| Total | 6980/100% |

In each group, the number of cases with the Sylvian fissure clearly shown only in the trans-cerebellar section were 1983, 1722, and 854 respectively, with clearness rates of 62.3%, 70.1%, and 63.6% respectively. The number of cases with the Sylvian fissure clearly shown only in the trans-thalamic section were 354, 258, and 122 respectively, with clearness rates of 11.1%, 10.5%, and 9.1% respectively. The number of cases with the Sylvian fissure clearly shown both in the trans-cerebellar and trans-thalamic section were 702, 368, and 174 respectively, with clearness rates of 22.1%, 15.0%, and 13.0% respectively. The number of cases with the Sylvian fissure unclearly shown both in the trans-cerebellar and trans-thalamic section were 143, 108, and 92 respectively, with unclearness rates of 4.5%, 4.4%, and 14.3% respectively. The clearness rate of showing Sylvian fissure on trans-cerebellar section was significantly higher than on trans-thalamic section (P < 0.05). The total clearness rate of showing Sylvian fissure on trans-cerebellar section and trans-thalamic section were 83.1% and 28.3% respectively. The data are shown in Table 2 and Table 3.

**Table 2.** Sample size and clearness rate of showing Sylvian fissure on different sections in different gestational weeks (n/%)

| Gestational weeks | sample size and clearness rate( n/%) |  |  |  |
| --- | --- | --- | --- | --- |
|  | only trans-cerebellar section clear | only trans-thalamic section clear | both trans-cerebellar and trans-thalamic section clear | both trans-cerebellar and trans-thalamic section unclear |
| 21~24 weeks | 1983/62.3% | 354/11.1% | 702/22.1% | 143/4.5% |
| 25~28 weeks | 1722/70.1% | 258/10.5% | 368/15.0% | 108/4.4% |
| 29~32 weeks | 854/63.6% | 122/9.1% | 174/13.0% | 192/14.3% |

**Table 3.**
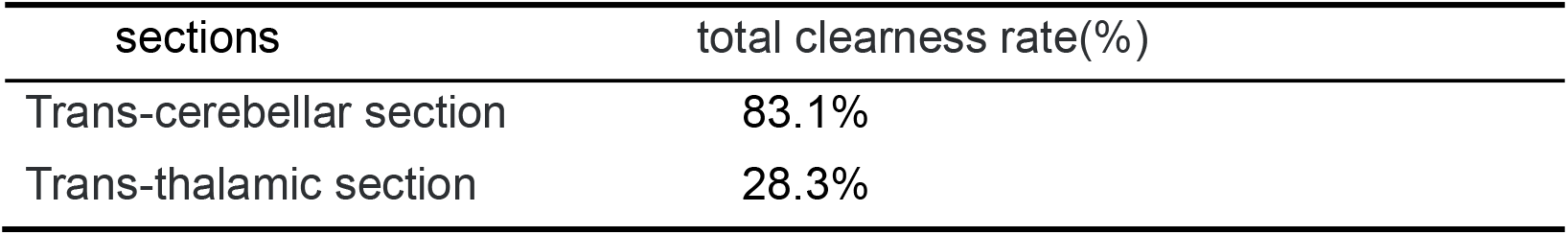
Comparison of total clearness rate of Sylvian fissure shown on different sections (%)

| sections | total clearness rate(%) |
| --- | --- |
| Trans-cerebellar section | 83.1% |
| Trans-thalamic section | 28.3% |

## DISCUSSION

In the process of brain development, due to the unbalanced development rate of intracranial cerebral parenchyma and skull, the increase of the whole surface area during the process of brain parenchyma development is faster than that of skull, and the development rate of the whole intracranial parenchyma is also unbalanced, which leads to the cerebral cortex invagination and folding to form different cerebral sulci^9^. The Sylvian fissure is one of the main cerebral sulcus, and its development is representative. Therefore, the development of the Sylvian fissure can be evaluated to further reveal the development law of the cerebral cortex. At present, there have been studies to evaluate the development law of the Sylvian fissure, but the research sections are not uniform, mainly including trans-thalamic section and custom section, and the research results are quite different, so there are no research results for promotion and application at present. In our preliminary clinical work, we found that the transverse section of the cerebellum showed the Sylvian fissure more clearly than the transverse section of the thalamus (as shown in Figure 1). In theory, the ultrasonic beam on the transverse section of the cerebellum was more perpendicular to the Sylvian fissure than that on the transverse section of the thalamus, so the transverse section of the cerebellum should also show the Sylvian fissure more clearly. To further confirm our hypothesis, in this study, we retrospectively studied the clearness rate of fetal Sylvian fissure on the transverse cerebellar and trans-thalamic sections.

**Figure 1.**
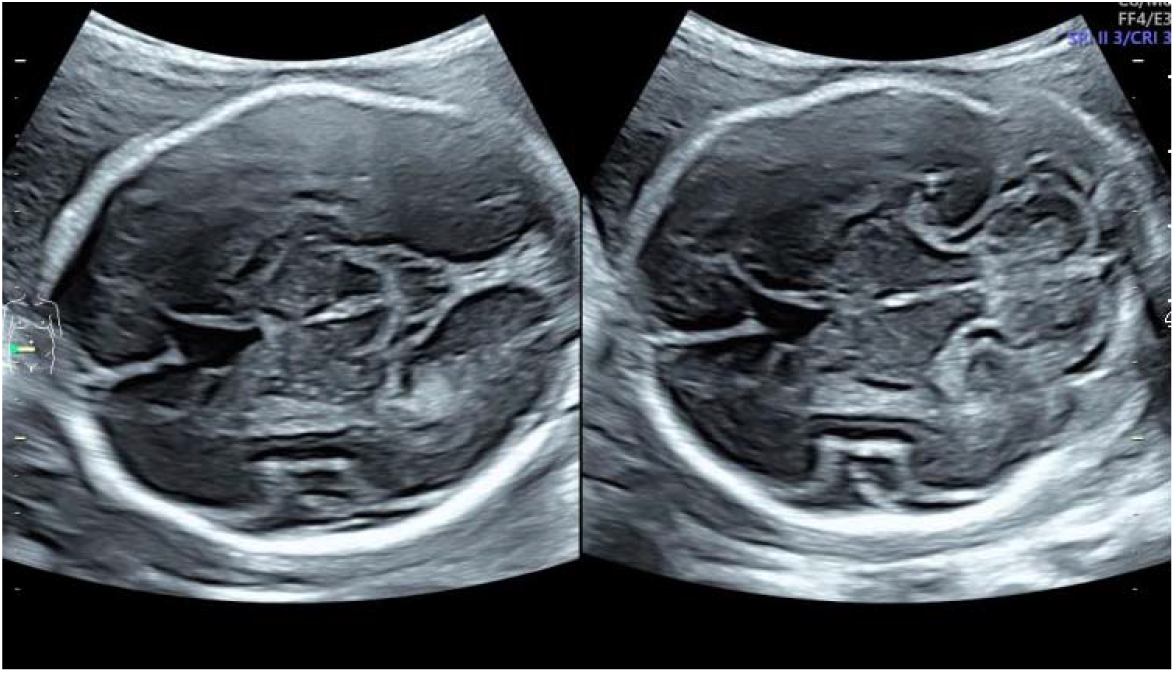
trans-thalamic and trans-cerebellar sections shown the fetal Sylvian fissure

It has been reported that 20 to 32 weeks of gestation is the occurrence time of the main cerebral gyrus^10^, so the subjects of this study were mainly included in the normal fetuses at 21 to 32 weeks of gestation, which were divided into three groups: 21 to 24 weeks, 25 to 28 weeks and 29 to 32 weeks of gestation. By dividing the gestational age into different groups, the clearness rate of Sylvian fissure in different sections of each group can be more comprehensively evaluated. At present, there is no reference standard for the assessment of the clearness rate of the Sylvian fissure of the fetus. In this study, the evaluation criteria for the clear appearance of the fetal Sylvian fissure was defined as follows: the morphology of the Sylvian fissure was complete, and the top and anterior edges of the Sylvian fissure were clear. In this study, the evaluation results of the customized Sylvian fissure were divided into four groups: only on trans-cerebellar section shown clear, only on trans-thalamic section shown clear, both on trans-cerebellar section and trans-thalamic section shown clear, and both on trans-cerebellar section and trans-thalamic section not shown clear. The results of this study showed that the clearness rate of the Sylvian fissure through the trans-cerebellar section was significantly higher than that through the trans-thalamic section during the 21 to 32 weeks of gestation in each group. The chi-square test was used to compare the clearness rate of the Sylvian fissure of the fetus, and the results were statistically different. In this study, the total clearness rate of Sylvian fissure was as high as 83% on the trans-cerebellar section, indicating that most fetuses could measure the developmental parameters related to the Sylvian fissure on this standard section, and the repeatability of the measured results would be good. The overall clearness rate of the Sylvian fissure through the trans-thalamic section was only 28%, indicating that in most cases, adjustments were needed on the standard section to measure the parameters related to the Sylvian fissure, and the repeatability of the measurement results would be affected.

In this study, a large sample size was retrospectively analyzed and it was concluded that prenatal ultrasound selection via trans-cerebellar section was superior to trans-thalamic section in evaluating developmental parameters related to the Sylvian fissure of the fetus.

## Data Availability

Data availability. The full data that supports the findings of this study are available from the corresponding author upon reasonable request.

## ACKNOWLEDGEMENTS

The authors thank all the women who participated in the study and acknowledge their significant contribution. Thank you to Zhang Dirong and Shi Yu for helping to design the research methods, Kong Fengbei and Yao Chunxiao for support with statistical analysis, She Ying and Wu Guoru for proposing some strategies for writing.

## Notes

### Competing Interest Statement

The authors have declared no competing interest.

### Funding Statement

Construction of medical key disciplines in Shenzhen city, Grant/Award Number: SZXK051; Shenzhen Virtual Reality Clinical Application Public Service Platform enhancement project, Grant/Award Number: XMHT20190104001; Three Projects Funded Project in Shenzhen city, Grant/Award Number: SZSM201512026

### Author Declarations

This study was approved by the Peking University Shenzhen Hospital and Health Service Human Research Ethics Committee. All pregnant women gave informed consent to this study.

